# Automated Neuroprognostication via Machine Learning in Neonates with Hypoxic-Ischemic Encephalopathy

**DOI:** 10.1101/2024.05.07.24306996

**Authors:** John D. Lewis, Atiyeh A. Miran, Michelle Stoopler, Helen M. Branson, Ashley Danguecan, Krishna Raghu, Linh G. Ly, Mehmet N. Cizmeci, Brian T. Kalish

## Abstract

**Objectives:** Neonatal hypoxic-ischemic encephalopathy is a serious neurologic condition associated with death or neurodevelopmental impairments. Magnetic resonance imaging (MRI) is routinely used for neuroprognostication, but there is substantial subjectivity and uncertainty about neurodevelopmental outcome prediction. We sought to develop an objective and automated approach for the analysis of newborn brain MRI to improve the accuracy of prognostication.

**Methods:** We created an anatomical MRI template from a sample of 286 infants treated with therapeutic hypothermia, and labeled the deep gray-matter structures. We extracted quantitative information, including shape-related information, and information represented by complex patterns (radiomic measures), from each of these structures in all infants. We then trained an elastic net model to use either only these measures, only the infants’ clinical data, or both, to predict neurodevelopmental outcomes, as measured by the Bayley Scales of Infant and Toddler Development at 18 months of age.

**Results:** Amongst those infants who survived and for whom Bayley scores were available for cognitive, language, and motor outcomes, we found sets of MRI-based measures that could predict their Bayley scores with correlations that were more than twice the correlations based on only the clinical data, and explained more than four times the variance in the observed scores; predictions based on the combination of the clinical and MRI-based measures were similar or marginally better.

**Interpretation:** Our findings show that machine learning models using MRI-based measures can predict neurodevelopmental outcomes in neonates with hypoxic-ischemic encephalopathy across all neurodevelopmental domains and across the full spectrum of outcomes.

## Introduction

Perinatal hypoxic-ischemic encephalopathy (HIE) affects approximately 1.5 infants per every 1000 births worldwide and is a major cause of death and neurodevelopmental disability (***Shankaran, 2015***). HIE is caused by a disruption in oxygen-rich blood flow to the fetus or neonate in the perinatal period. The implementation of therapeutic hypothermia in neonates with HIE has improved outcomes, but still, nearly half of infants with HIE die or develop neurodevelopmental impairments, including cerebral palsy, cognitive delay, speech and language problems, and behavioral disorders. (***Simbruner et al., 2010; Azzopardi et al., 2014; Cheong et al., 2012; Groenendaal et al., 2013; Glass, 2018; Nair and Ku-mar, 2018; Finder et al., 2020; Schreglmann et al., 2020; Steinmetz et al., 2024***). Today, brain magnetic resonance imaging (MRI) is routinely used for supporting the diagnosis of HIE and also for neuroprognostication in this population (***Martinez-Biarge et al., 2010; Rutherford et al., 2010; Cheong et al., 2012; Shankaran et al., 2012; Li et al., 2013; Hayes et al., 2016; Alderliesten et al., 2017; Weeke et al., 2018; Aker et al., 2022***). In particular, injuries to the deep gray matter (DGM), posterior limb of the internal capsule (PLIC), cerebral peduncles, cortex, and watershed zones have been associated with neurodevelopmental impairment after HIE (***Martinez-Biarge et al., 2010; Rutherford et al., 2010; Hayes et al., 2016; Weeke et al., 2018; Ouwehand et al., 2020; Aker et al., 2022***). However, the interpretation of neonatal brain MRI relies upon extensive neuroradiology expertise, is time-intensive, and subject to inter-rater variability.

Recent developments in radiomics provide a means to quantify brain injury more precisely, and machine learning models can utilize these new neuroimaging measures together with clinical and laboratory parameters to form a more objective prognosis. In this study, we utilized this approach to predict neurodevelopmental outcomes in a single institution cohort of neonates with HIE. We hypothesized that this approach would enhance the accuracy of prognostication.

## Materials and Methods

### Study Cohort, Clinical and Laboratory Parameters

This retrospective cohort study was conducted at the Hospital for Sick Children in Toronto, Canada. The Institutional Research Ethics Board reviewed and approved the study protocols (REB:1000064940,1000079302) and waived informed consent. Infants with a gestational age of >35 weeks with perinatal HIE who underwent therapeutic hypothermia between January 2018 and January 2022 were included. HIE was diagnosed based on the presence of neonatal encephalopathy in infants with presumed perinatal asphyxia and defined by one or more of the following: Apgar score of ≤5 at 10 minutes, cord arterial blood or first-hour blood gas pH <7.0 or base deficit ≥16 mmol/L, or the need for resuscitation at birth. The severity of HIE was classified clinically according to the Sarnat score (Sarnat et al., 1976) and Thompson score (Thompson et al. 1997).

Therapeutic hypothermia was initiated within 6 hours after birth and continued for 72 hours, as per institutional protocols, unless, in rare circumstances, discontinued early due to clinical contraindications. The target core temperature was maintained at 33° to 34° Celsius with the whole-body cooling system. Infants were excluded for major congenital anomalies, chromosomal or genetic abnormalities, or neonatal encephalopathy due to causes other than HIE. Clinical data were obtained from the electronic hospital database. Gestational age, birth weight, sex, 5-minute Apgar score, umbilical cord arterial and venous pH, first postnatal gas pH, highest blood lactate within the first 72 hours, and highest Thompson score were used. The details of these parameters are provided in Table 1. As can be seen in Table 1, there are missing values for many of these measures ; this missing data was dealt with using imputation. We used the MICE Imputation method in scikit-learn (***Pedregosa et al., 2011***). MICE, short for ‘Multiple Imputation by Chained Equation’ is an advanced missing data imputation technique that uses multiple iterations of a Machine Learning model trained to predict the missing values in the data using the known values as predictors (***Van Buuren and Oudshoorn, 2000; Van Buuren and Groothuis-Oudshoorn, 2011***). We used 10 iterations for each analysis that included variables which had missing values. Additionally, for one infant with severe CP and global developmental delay for whom the Bayley-III assessment was not feasible, a percentile score of 1 was assigned across all domains.

**Table 1.** The demographic and clinical data for the study population.

| | Female (111)<br>$\mu \pm \sigma (N)$ | Male (175)<br>$\mu \pm \sigma (N)$ | *p*-value of diff. |
| --- | --- | --- | --- |
| **Gestational age, weeks** | 38.99 ± 1.65 (111) | 38.64 ± 1.87 (175) | 0.10 |
| **Birth weight, g** | 3, 208.38 ± 548.32 (111) | 3, 240.15 ± 635.93 (175) | 0.65 |
| **5-minute Apgar score** | 4.44 ± 2.12 (103) | 4.55 ± 2.26 (168) | 0.68 |
| **Cord venous pH** | 7.10 ± 0.16 (97) | 7.05 ± 0.52 (152) | 0.20 |
| **Cord arterial pH** | 7.00 ± 0.15 (98) | 7.05 ± 0.66 (153) | 0.39 |
| **First postnatal pH** | 7.10 ± 0.17 (105) | 7.12 ± 0.42 (157) | 0.61 |
| **Highest lactate, mmol/l** | 9.10 ± 4.96 (110) | 9.36 ± 5.21 (175) | 0.67 |
| **Highest Thompson score** | 8.99 ± 4.09 (94) | 8.64 ± 3.38 (145) | 0.49 |

### MRI Acquisition and Processing

All neonates (n=357) underwent brain MRI as near as possible to 4 days after birth, after the completion of therapeutic hypothermia. The MRI scans were acquired on a 3 Tesla scanner (Magnetom Skyra, Siemens Healthcare Limited, Germany) or a 1.5 Tesla scanner (Ingenia, Philips NV, Netherlands) with an age-appropriate head coil. The acquisition protocol on both scanners produced high-resolution 3D T1-weighted volume(s), and 2D T2-weighted volumes with high in-plane resolution, but thick slices, in each of axial, coronal, and sagittal orientations. The 3D T1-weighted images were acquired as sagittal slices, with a slice-thickness of 0.5 mm, and an in-plane resolution of 0.4018 mm x 0.4018 mm (echo time 3.52 ms ; repetition time 2200 ms). The 2D T2-weighted volumes were acquired with a slice thickness of 3 mm, an in-plane resolution of 0.5 mm x 0.5 mm, and with 3.3 mm spacing between slices (echo time 186 ms ; repetition time 5330 ms). In an attempt to obtain usable data, the scan operator might collect multiple T1- and T2-weighted images. We performed quality control on the data, and eliminated data with artifacts, *e.g*. motion. There were 286 infants with acceptable T1- and T2-weighted data ; 220 infants acquired on the Siemens scanner, and 66 infants acquired on the Philips scanner. The demographic, clinical and laboratory data for these infants are presented in Table 1.

All acceptable T1-weighted volumes were then denoised with DenoiseImage (Manjón et al., 2010b), and non-uniformity corrected with N4BiasFieldCorrection (Nicholas et al., 2010). If there were multiple acceptable T1-weighted volumes for a subject, one was chosen, and the others were aligned to it with a rigid registration using ANTs (Avants et al. 2009). All aligned volumes were then averaged, resampled to 0.5 mm iso, and normalized to have intensity values between 0 and 100.

The T2-weighted images were processed with super-resolution code (Manjón et al., 2010a) to produce 0.5 mm iso volumes. Each such volume was then denoised, non-uniformity corrected, normalized to have intensities between 0 and 100, and then linearly registered to its T1-weighted counterpart using ANTs. All T2-weighted volumes, in all three orientations were then averaged.

A brain mask was extracted by providing the T1- and T2-weighted volumes to a convolutional neural network (CNN) trained to do this. The training data for the CNN was progressively constructed by registering the FinnBrain neonate multi-contrast template (Tuulari et al., 2024) to each subject’s T1- and T2-weighted volumes, using the resulting transform to bring the mask from the Finnbrain template to the subject, and then, if the result was approximately correct, manually correcting the result and adding it to the training set. The resulting brain masking tool can be found at https://gin.g-node.org/johndlewis/HIE/Tools/BET-CNN.sh.

### Template Construction and Use

Once we had masked, denoised, non-uniformity corrected, T1- and T2-weighted volumes for each subject, we then provided these data to the ANTs script *antsMultivariateTemplateConstruction2.sh*, which we ran in four stages. First, we ran it with the FinnBrain neonate T1- and T2-weighted template as a target, and used rigid registration to build a population-specific target for our data. Second, we ran affine registration, starting from the template arrived at via rigid registration. Third, we ran the non-linear SyN registration method with the result of the second stage as the target. This new population specific neonatal brain multi-contrast template was then linearly and nonlinearly registered to the multi-contrast neonatal template from the FinnBrain Birth Cohort Study (Tuulari et al., 2024) using ANTs. The inverse of the resulting transform was then used to overlay the labels from the FinnBrain neonatal template on our new population specific neonatal brain template. Our final template is shown in Figure 1, with the T1-weighted volume shown on the top, the T2-weighted volume below, and the T2-weighted volume with the labels overlain on the bottom. These labels, as well as labels for the left and right PLIC, were overlaid on individual subjects by linearly and nonlinearly registering the subject data to the template, then using the inverse of the resulting transform to take the template labels back to the subject. Once the labels were on a subject, the geometric measures were taken by running *LabelGeometryMeasures* and the radiomic features of both the T1- and T2-weighted volumes for each structure were taken by running *pyRadiomics*, with each label as a mask (***Gillies et al., 2016; Wagner et al., 2021, 2022***); this was done separately for both hemispheres.

**Figure 1.**
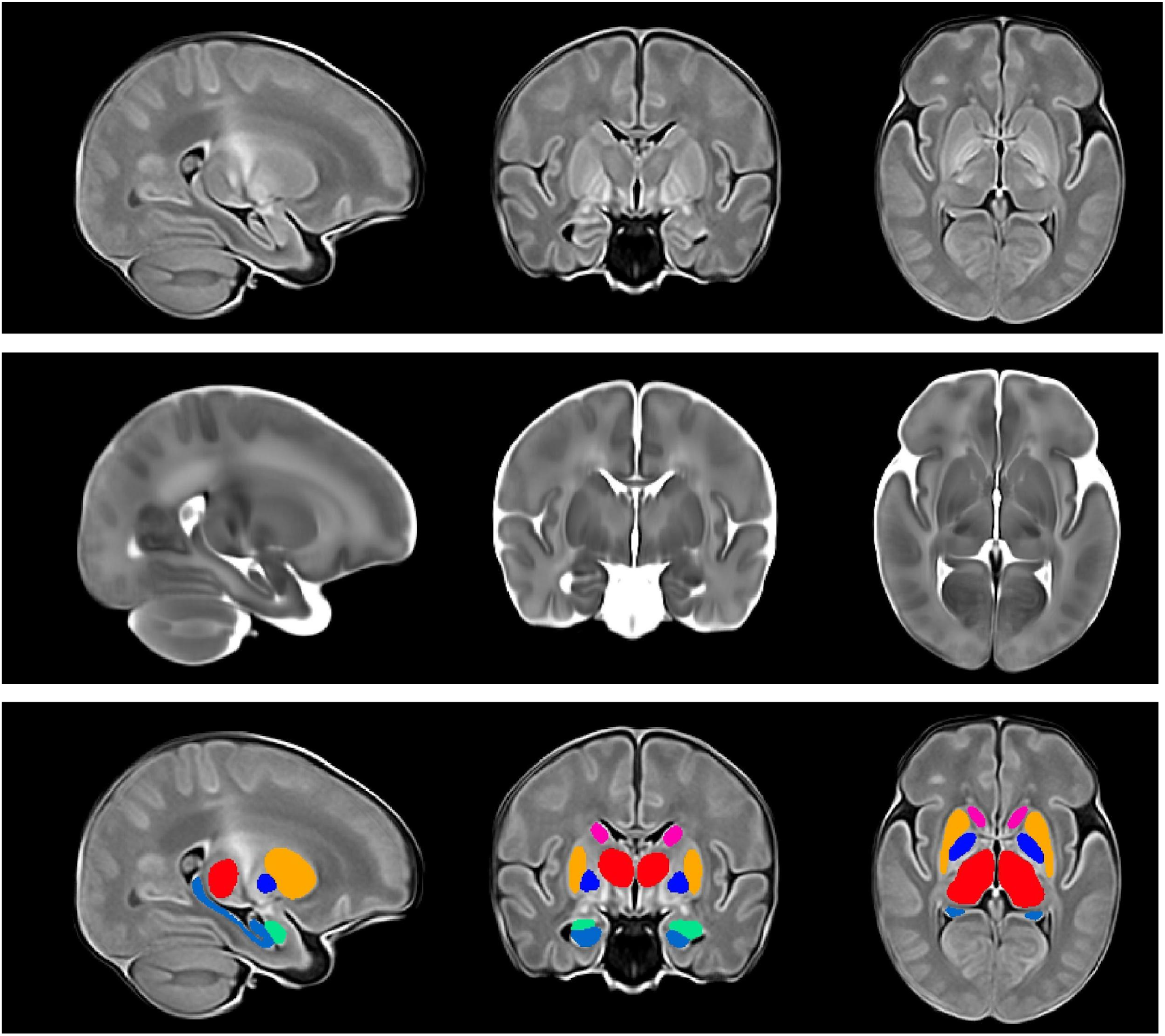
The new population specific neonatal brain multi-contrast template. The top row shows the T1-weighted volume ; the second row shows the T2-weighted volume ; the bottom row shows the t2-weighted volume with the labels for the amygdala, hippocampus, and subcortical gray structures overlaid on it. The amygdala is shown in green ; the hippocampus in light blue ; the globus pallidus in dark blue ; the putamen in gold ; the caudate in pink ; and the thalamus in red.

Radiomics capture complex patterns that may fail to be seen with the naked eye (***Yip et al., 2017***), including features of the image intensity histogram; the relationships between image voxels; neighborhood gray-tone difference derived textures, and features of complex patterns. Descriptions of each of the radiomics measures can be found in the pyradiomics documentation!.

### Analysis of Relation Between Measures and Outcomes

Gestational age, sex, and the clinical parameters, or the MRI-based measures, or the combination, were input to a linear regression model to predict 18-month neurodevelopmental outcomes. The regression model was used to predict seven different outcome scores on the Bayley Scales of Infant and Toddler Development, 3rd edition (Bayley-III) at 18 months corrected age: cognitive, receptive language, expressive language, composite language, gross motor, fine motor, and composite motor.

The linear regression model utilized Elastic-Net penalized linear regression. Elastic-Net is designed to balance two approaches to regularization of the coefficients: the approach used in Lasso regression, and the approach used in Ridge regression (Zou and Hastie, 2005). Ridge regression adds the sum of the squares of the coefficients to the sum of squares of the residuals. That keeps the coefficients small, but keeps all variables in the model. Lasso regression adds the sum of the absolute value of the coefficients. That allows some coefficients to go to zero ; thus some features of the data may be ignored. Elastic-Net aims for a balance which allows for learning a sparse model where few of the weights are non-zero, and the coefficients are generally kept from becoming large. This balance is controlled by hyper-parameters that determine the size of the penalties that are incurred, and the weighting between the choices. We used 10-fold cross-validation to ensure that our results generalize; and within each fold we used 10-fold cross-validation to find the best hyperparameters. For each outer fold, the elastic-net model is fitted on the training data, and predictions made for the testing data, *i.e*. data that the model has not been fitted for. To quantify the performance of the linear regression model, we used three evaluation metrics: the correlation coefficient (R) between the predicted and observed outcomes; the coefficient of determination (*R*2), which is an estimate of the proportion of variance in the observed outcomes that can be explained by the predictors; and the mean absolute error (MAE) of the predicted outcomes versus the measured outcomes.

We first assessed the predictions based on only the clinical data. We then assessed the predictions based on the MRI-based measures. Lastly, we assessed the combination of the clinical and the MRI-based measures. The models were further analyzed to determine which aspects of the inputs were driving the predictions.

## Results

The analyses used the data from all of the neonates that survived, and for whom we had post-rewarming brain MRI, and Bayley-III scores. For the infants for whom we had Bayley-III cognitive outcome scores (n=174), brain MRI measures yielded a correlation coefficient for the cognitive outcomes that is more than twice that of the correlation produced with the clinical measures alone (r:0.484, 95% CI[0.361, 0.590] vs. r:0.225, 95% CI [0.079, 0.362], respectively) and the relation based on the brain MRI data explained more than four times the variance in the observed outcome compared to that of the relation based on the clinical measures (r^2^:0.234 vs 0.050, respectively). Combining clinical and MRI metrics did not further improve the predictive accuracy (r:0.452, 95% CI [0.325, 0.563]). Measures from each structure contributed to the results from the analyses which used the MRI-based measures; in the analysis which used the combined measures, only birth weight contributed to the result. The largest contribution to the result came from the PLIC, and 10.6% of the predictors came from radiomic features from the PLIC. But a number of predictors came from the hippocampus (14.9%), amygdala (13.8%), thalamus (8.5%), and caudate (6.4%); and radiomic features of the brain as a whole constituted 40.4% of the cognitive outcome predictors. Among the clinical and laboratory parameters, only birth weight was selected by the model. The contribution of the top predictors of cognitive outcome for the combined model is presented in Figure 2. The contributions of the full set of predictors for each result can be found in the supplementary material.

**Figure 2.**
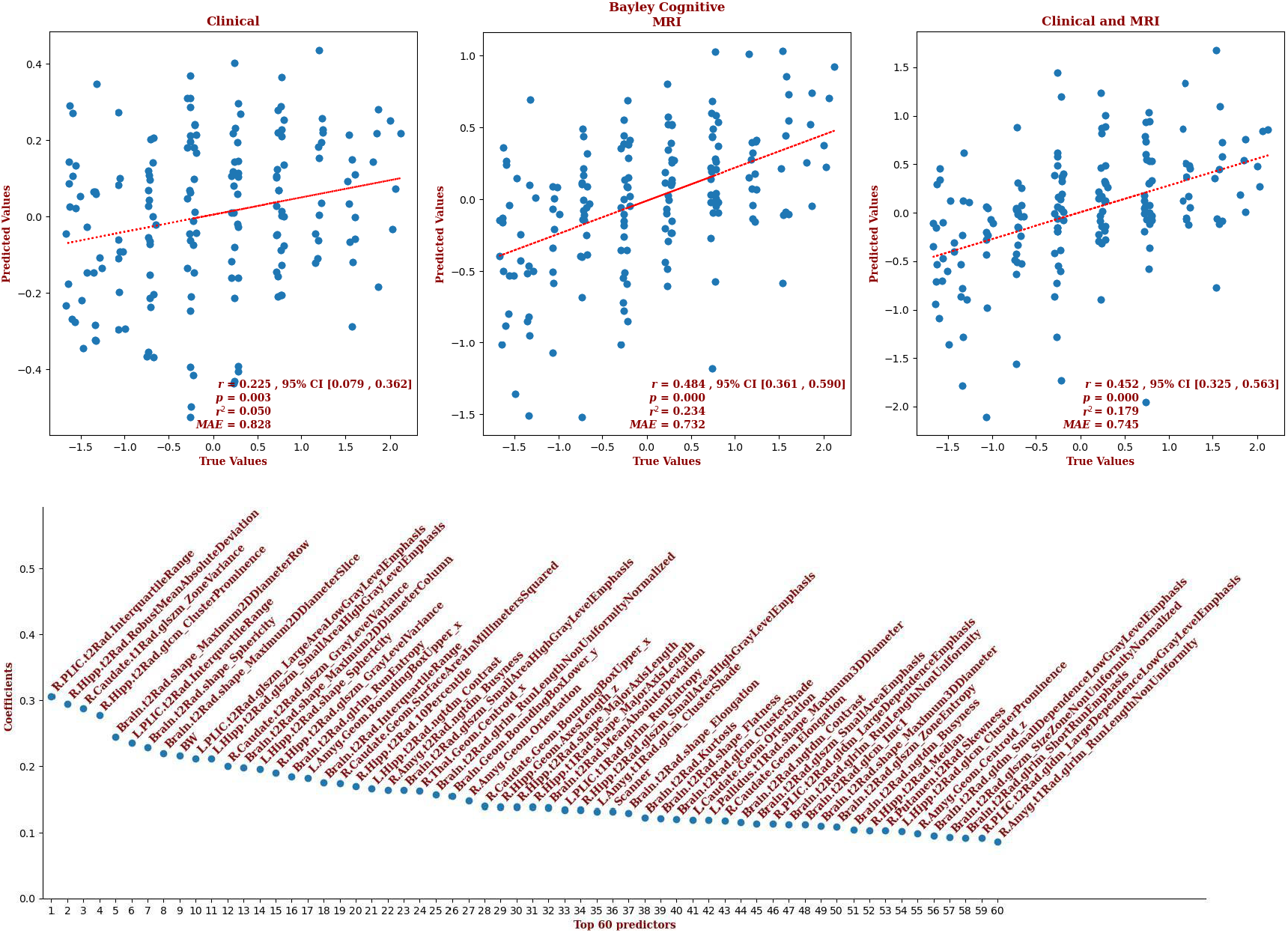
The regression results for the Bayley cognitive scores using (left) only the clinical variables ; (center) only the MRI measures ; and (right) both the clinical and MRI measures. Note that the correlation based on the MRI measures is more than twice that of the correlation based on the clinical measures, and accounts for more than four times the variance in the data ; the correlation based on the combined measures is approximately the same as that based on the MRI measures alone. The top predictors for the result based on the combined measures are shown at the bottom, with the magnitude of their coefficients on the y-axis. Note that the radiomic features include first order statistics, neighbourhood gray tone difference metrics (ngtdm), gray level size zone metrics (glszm), gray level run length metrics (glrlm), gray level co-occurence metrics (glcm), and shape metrics.

For the infants for whom we had Bayley-III expressive language outcomes (n=153), brain MRI measures showed a correlation more than three times that of the correlation based on the clinical measures (r:0.509, 95% CI [0.381, 0.618] vs. r:0.152, 95% CI [-0.007, 0.303], respectively) and explained more than eleven times the variance in the observed outcome (r^2^:0.253 vs 0.022, respectively). Combining clinical and MRI metrics did not further improve the predictive accuracy (r:0.500, 95% CI [0.371, 0.610]). The largest contributions to the result came from the amygdala, and 26.3% of the predictors came from features from the amygdala. But features of the thalamus, putamen, and PLIC each comprised 15.8% of the predictors; features of the globus pallidus and caudate comprised 10.5% and 5.3% of the predictors, respectively. The only clinical metric that was selected by the model was gestational age. The contribution of the predictors of expressive language for the combined model is presented in Figure 3. The contributions of the full set of predictors for each result can be found in the supplementary material.

**Figure 3.**
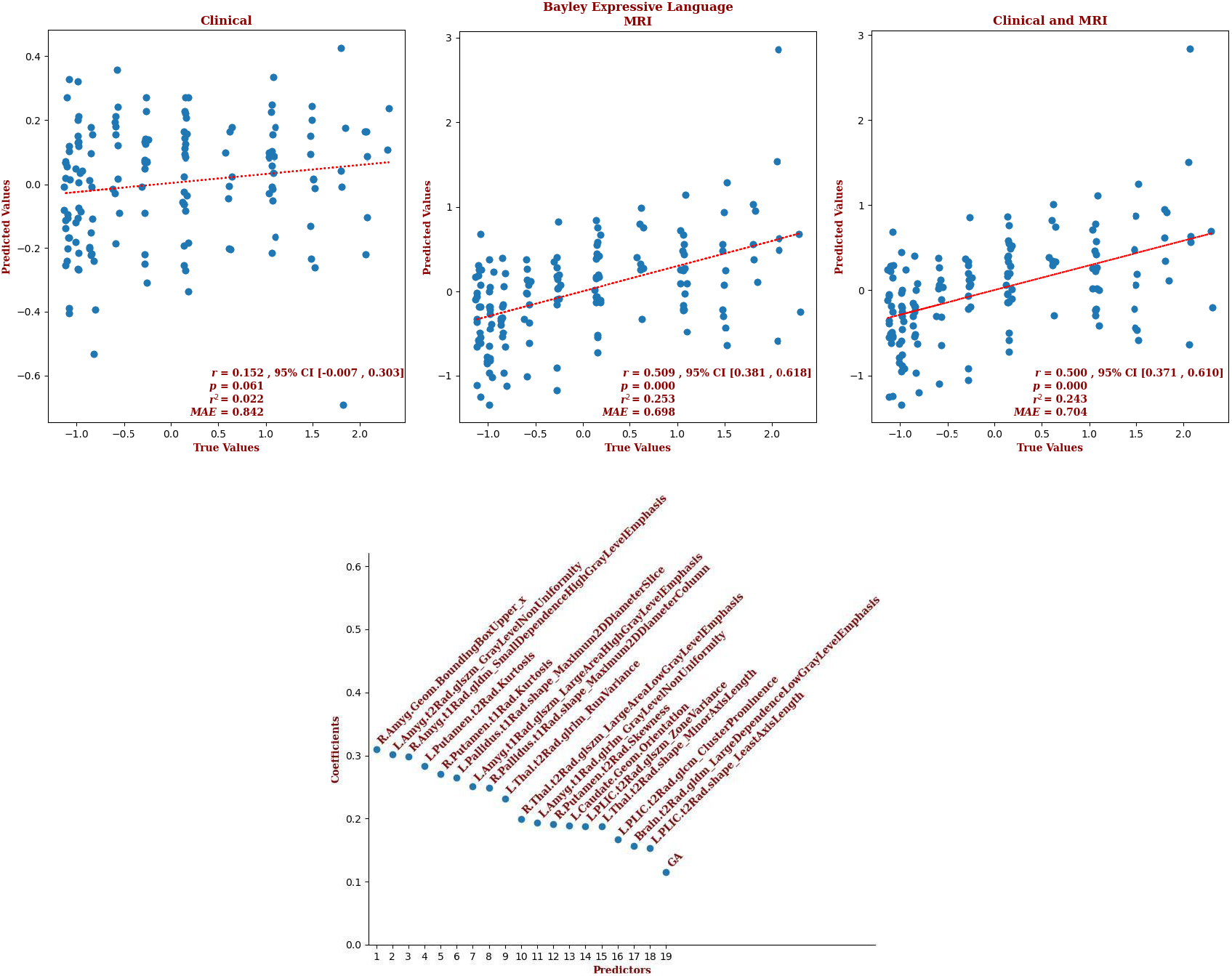
The regression results for the Bayley expressive language scores using (left) only the clinical variables ; (center) only the MRI measures ; and (right) both the clinical and MRI measures. Note that the correlation based on the MRI measures is more than three times that of the correlation based on the clinical measures, and accounts for eleven and a half times the variance in the data; the correlation based on the combined measures is approximately the same as that based on the MRI measures alone. Note also, that the correlation based on the clinical measures is only marginally significant. The predictors for the result based on the combined measures are shown at the bottom, with the magnitude of their coefficients on the y-axis. That these are the only measures used by the model is notable, as well as the prominence of the amygdala amongst the predictors. Note that the radiomic features include first order statistics, gray level dependency metrics (gldm), gray level size zone metrics (glszm), gray level run length metrics (glrlm), gray level co-occurence metrics (glcm), and shape metrics.

For the infants for whom we had Bayley-III receptive language outcomes (n=153), brain MRI measures yielded a correlation more than twice that of the correlation based on the clinical measures (r:0.554, 95% CI [0.433, 0.655] vs. r:0.244, 95% CI [0.089, 0.388], respectively) and explained more than five times the variance in the observed outcome data (r^2^:0.307 vs 0.055, respectively). Combining clinical and MRI metrics did not further improve the predictive accuracy (r:0.558, 95% CI [0.438, 0.658]). The largest contribution to the result came from the PLIC, and 7.4% of the predictors came from the geometric and radiomic features from the PLIC. But features of each brain structure contributed to the predictions of the combined model ; features of the amygdala made up 13.9% of the predictor set, followed by features of the putamen (12%), the caudate (9.3%), the hippocampus and PLIC (7.4% each), the thalamus (6.5%), the globus pallidus (1.9%), and the brain as a whole (39.8%). The two clinical predictors that were selected by the model were gestational age and sex ; and it is notable that sex is prominent, being the second largest contributor to the result. The contribution of the top predictors of receptive language for the combined model is presented in Figure 4. The contributions of the full set of predictors for each result can be found in the supplementary material.

**Figure 4.**
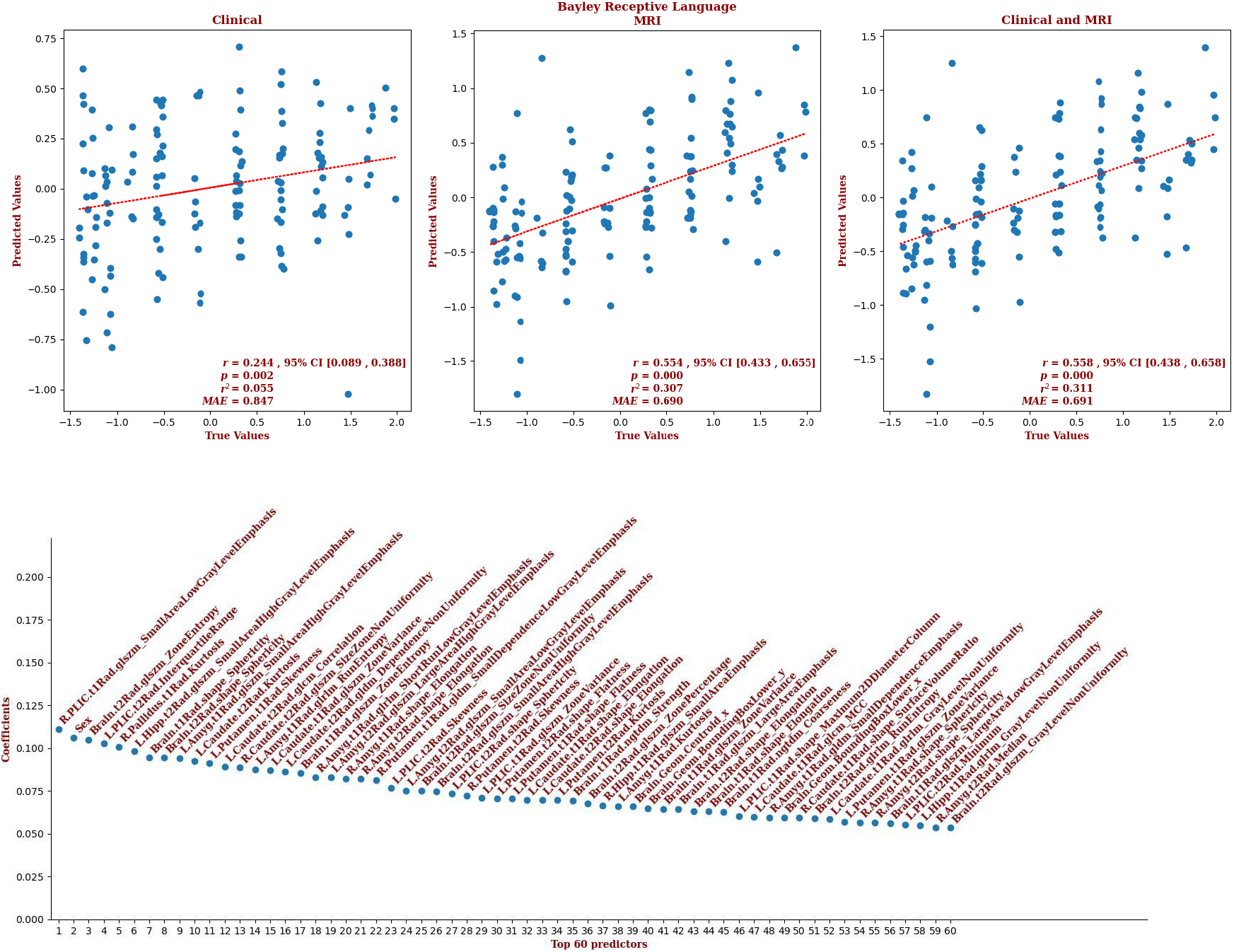
The regression results for the Bayley receptive language scores using (left) only the clinical variables ; (center) only the MRI measures ; and (right) both sets of measures together. Note that the correlation based on the MRI measures is more than twice that of the correlation based on the clinical measures, and accounts for more than five and a half times the variance in the data ; the correlation based on the combined measures is approximately the same as that for the MRI alone. The top predictors for the results based on the combined measures are shown at the bottom, with the magnitude of their coefficients on the y-axis. ; notably, the PLIC is a top predictor, as well as sex. Note that the radiomic features include first order statistics, neighbourhood gray tone difference metrics (ngtdm), gray level size zone metrics (glszm), gray level run length metrics (glrlm), gray level co-occurence metrics (glcm), and shape metrics.

For the infants for whom we had Bayley-III composite language outcomes (n=155), brain MRI measures showed a correlation more than twice that of the correlation based on the clinical measures (r:0.634, 95% CI [0.529, 0.720] vs. r:0.259, 95% CI [0.105, 0.400], respectively) and explained more than six times the variance in the observed outcomes (r^2^:0.396 vs 0.065, respectively). Combining clinical and MRI metrics marginally improved the predictive accuracy (r:0.668, 95% CI [0.564, 0.743]). The largest contribution to the result came from the PLIC, but only 4% of the predictors came from the PLIC. Most predictors came from the amygdala (34.7%), followed by the brain as a whole (32.7%), the putamen (8%), and the caudate and globus pallidus (6% each). Gestational age and sex were significant clinical predictors. The detailed contribution of each parameter to composite language outcome prediction is presented in Figure 5 and provided in the supplementary material.

**Figure 5.**
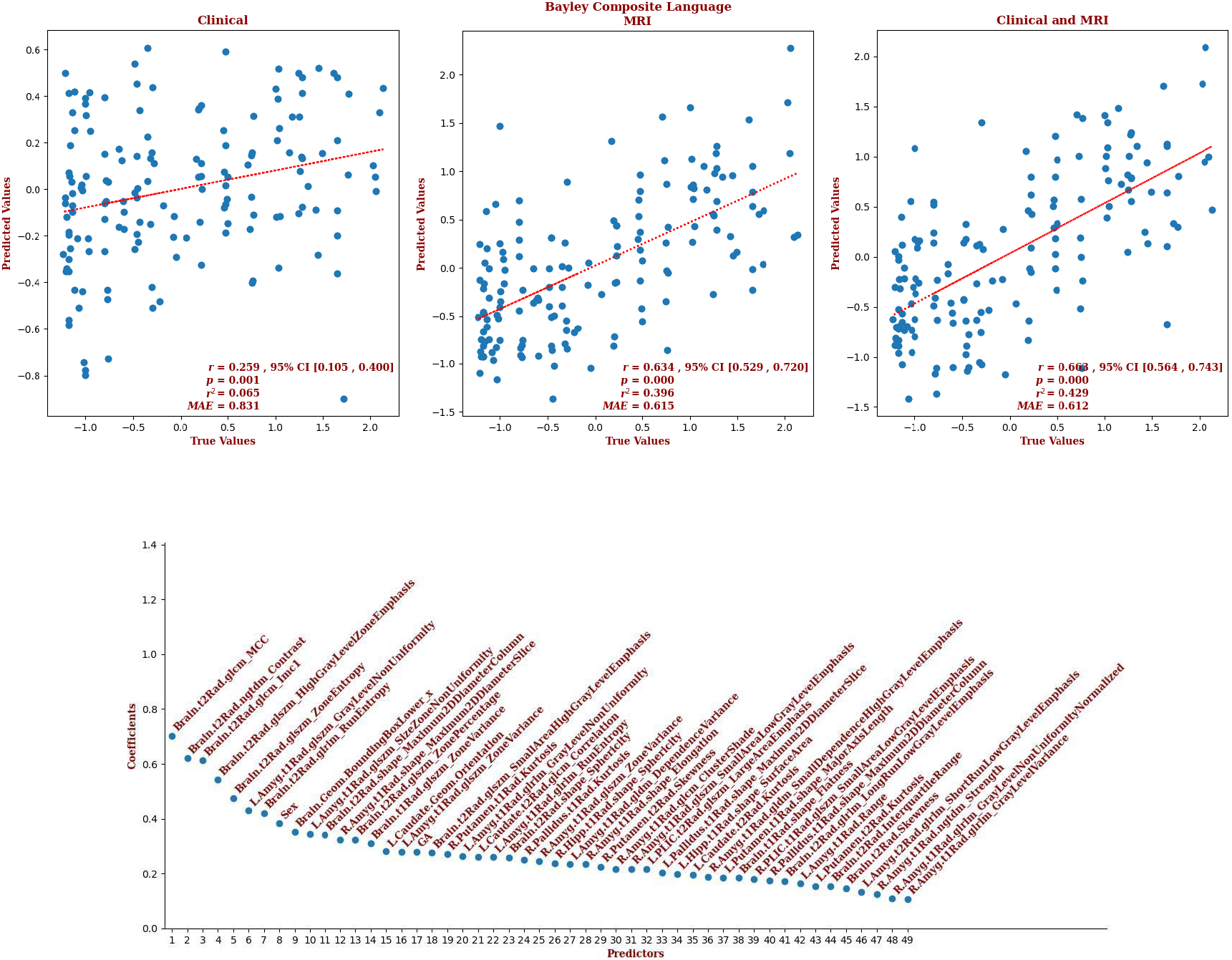
The regression results for the Bayley composite language scores using (left) only the clinical variables ; (center) only the MRI measures ; and (right) both sets of measures. Note that the correlation based on the MRI measures is almost two and a half times that of the correlation based on the clinical measures, and accounts for more than six times the variance in the data ; and the correlation based on the combination of measures is slightly better still. The predictors for the result based on the combined measures are shown at the bottom, with the magnitude of their coefficients on the y-axis. That this is the full set of predictors is notable, as well as the prominence of measures from the brain, i.e. not the deep-gray structures. Note that the radiomic features include first order statistics, neighbourhood gray tone difference metrics (ngtdm), gray level size zone metrics (glszm), gray level run length metrics (glrlm), gray level co-occurence metrics (glcm), and shape metrics.

For the infants for whom we had Bayley-III gross motor outcomes (n=161), brain MRI measures yielded a correlation almost three times that of the correlation based on the clinical measures (r:0.492, 95% CI [0.365, 0.601] vs. r:0.168, 95% CI [0.120, 0.215], respectively) and explained more than ten times the variance in the observed outcome data (r^2^:0.242 vs 0.024, respectively). Combining the clinical and MRI metrics marginally improved the predictive accuracy (r:0.510, 95% CI [0.473, 0.546]). The largest contribution to the result came from the caudate, and 15.2% of the predictors came from features of the caudate. But an equal number of predictors came from the brain as a whole, and the most predictors came from the PLIC (21.7%) ; the putamen and amygdala both contributed 10.9% of the predictors, followed by the globus pallidus (6.5%), the hippocampus (4.3%), and the thalamus (2.2%). Gestational age, sex, 5-minute Apgar score, the highest Thompson score, blood lactate, and venous pH were significant clinical predictors. The detailed contribution of each parameter to gross motor outcome prediction is presented in Figure 6 and provided in the supplementary material.

**Figure 6.**
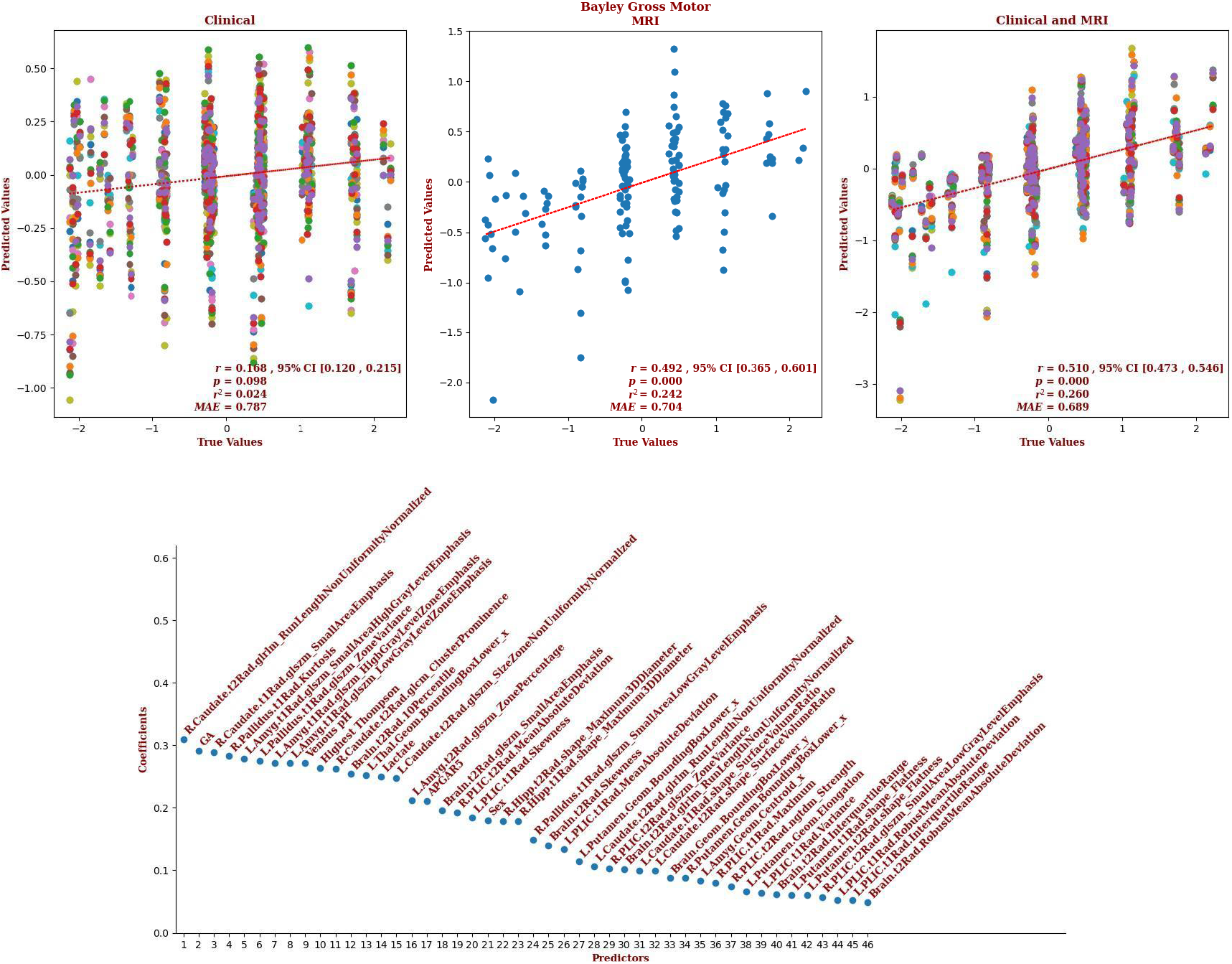
The regression results for the Bayley gross motor scores using (left) only the clinical measures ; (center) only the MRI measures ; and (right) the combined sets of measures. The multiple colors on the plots for the clinical measures and the combined measures indicate that the models retained variables for which there were missing values ; each color represents a different imputation. Note that the correlation based on the MRI measures is almost three times that of the correlation based on the clinical measures, and accounts for more than ten times the variance in the data ; the result for the combined sets of measures is slightly better still. Note also, that the correlation based on the clinical measures is only marginally significant. The predictors for the result based on the combined measures are shown at the bottom, with the magnitude of their coefficients on the y-axis. That this is the full set of predictors is notable, as well as the prominence of measures from the caudate and amgdala. Note that the radiomic features include first order statistics, neighbourhood gray tone difference metrics (ngtdm), gray level size zone metrics (glszm), gray level run length metrics (glrlm), gray level co-occurence metrics (glcm), and shape metrics.

For the infants for whom we had Bayley-III fine motor outcomes (n=165), brain MRI measures showed a correlation more than twice that of the correlation based on the clinical measures (r:0.597, 95% CI [0.489, 0.687] vs. r:0.292, 95% CI [0.247, 0.336], respectively) and explained more than four times the variance in the observed outcome data (r^2^:0.354 vs 0.083, respectively). Combining clinical and MRI metrics only marginally improved the predictive accuracy (r:0.609, 95% CI [0.577, 0.638]). The largest contribution to the result came from the caudate, and 15% of the predictors came from features of the caudate. But an equal number of predictors came from globus pallidus, and the most predictors came from the hippocampus (50%), with the brain as a whole providing another 10% of the predictors, and the putamen providing another 5%. The only clinical predictor retained by the model was venous pH. The detailed contribution of each parameter to fine motor outcome prediction is presented in Figure 7 and provided in the supplementary material.

**Figure 7.**
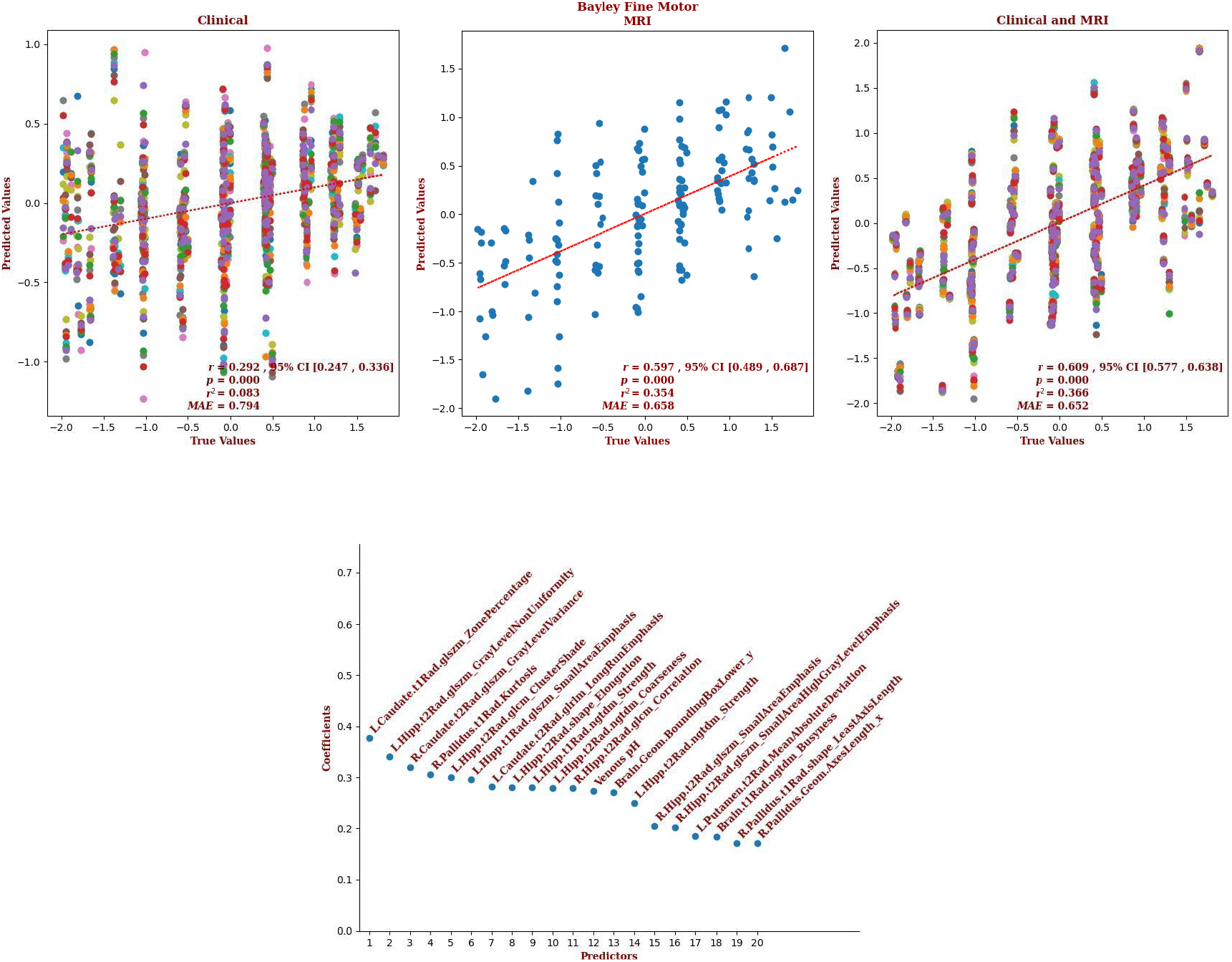
The regression results for the Bayley fine motor scores using (left) only the clinical measures ; (center) only the MRI measures ; and (right) the combined sets of measures. The multiple colors on the plot for the clinical measures indicate that the model retained variables for which there were missing values ; each color represents a different imputation. Note that the correlation based on the MRI measures is more than two times that of the correlation based on the clinical measures, and accounts for more than four times the variance in the data ; the result for the combined measures is slightly better still. The predictors for the result based on the combined measures are shown at the bottom, with the magnitude of their coefficients on the y-axis. That this is the full set of predictors is notable, as well as the prominence of measures from the caudate and the hippocampus. Note that the radiomic features include first order statistics, neighbourhood gray tone difference metrics (ngtdm), gray level size zone metrics (glszm), gray level run length metrics (glrlm), gray level co-occurence metrics (glcm), and shape metrics.

For the infants for whom we had Bayley-III composite motor outcomes (n=166), brain MRI measures showed a correlation more than twice that of the correlation based on the clinical measures (r:0.506, 95% CI [0.383, 0.611] vs. r:0.242, 95% CI [0.196, 0.287], respectively) and explained more than four times the variance in the observed outcome data (r^2^:0.255 vs 0.059, respectively). Combining clinical and MRI metrics marginally improved the predictive accuracy (r:0.512, 95% CI [0.476, 0.547]). The largest contribution to the result came from the caudate, and 11.8% of the predictors came from features of the caudate. But the largest number of predictors came from the hippocampus (29.4%), and the brain as a whole supplied 23.5% of the predictors. The globus pallidus, PLIC, and putamen each supplied 5.8% of the predictors. Gestational age and venous pH were the significant clinical predictors. The detailed contribution of each parameter to the composite motor outcome prediction is presented in Figure 8 and provided in the supplementary material.

**Figure 8.**
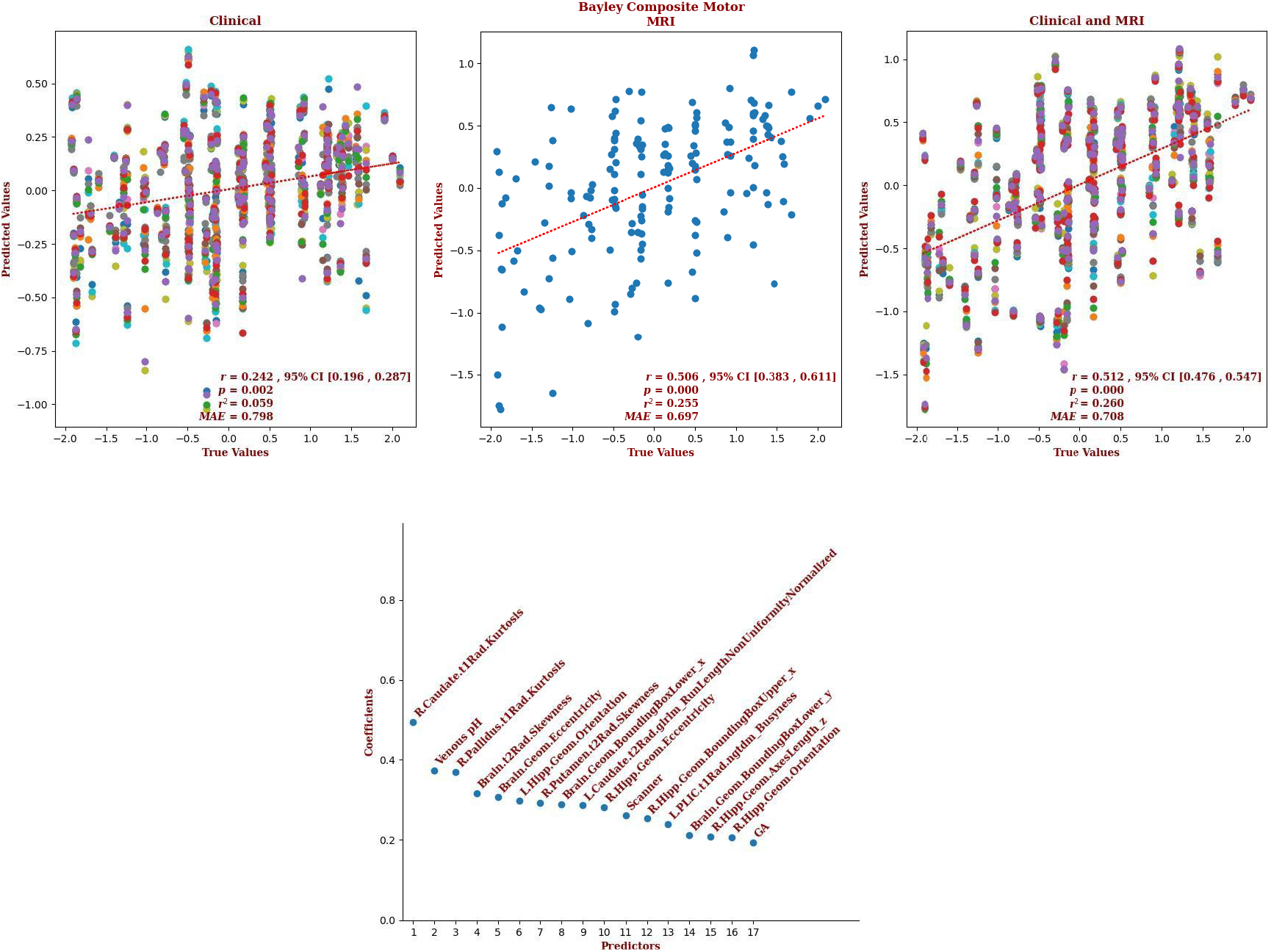
The regression results for the Bayley composite motor scores using (left) only the clinical measures ; (center) only the MRI measures ; and (right) the combined sets of measures. The multiple colors on the plot for the clinical and combined measures indicate that the model retained variables for which there were missing values ; each color represents a different imputation. Note that the correlation based on the MRI measures is more than two times that of the correlation based on the clinical measures, and accounts for almost four and a half times the variance in the data ; and the results for the combined measures is slightly better still. The predictors for the result based on the combined measures are shown at the bottom, with the magnitude of their coefficients on the y-axis. That this is the full set of predictors is notable, as well as the prominence of measures from the caudate, the pallidus, the putamen, the hippocampus, the PLIC, and the whole brain, suggesting wide-spread involvement. Note that the radiomic features include first order statistics, neighbourhood gray tone difference metrics (ngtdm), and gray level run length metrics (glrlm).

## Discussion

Post-rewarming MRI obtained in neonates with HIE is commonly used to assess the extent and severity of brain injury and counsel caregivers about the expected neurodevelopmental outcome trajectories. Although the implementation of hypoxic-ischemic injury scoring systems for neonatal MRI has improved this process, these scoring systems are not widely employed in the clinical setting and are generally reserved for research purposes, as they are time-consuming and require significant neonatal neuroimaging interpretation expertise (***Weeke et al., 2018; Bach et al., 2021; Machie et al., 2021; O’Kane et al., 2021; Bhroin et al., 2022***). Previous studies have shown that severe and extensive brain injury on MRI is predictive of mortality or severe neurodevelopmental disability; however, milder forms of brain injury have been claimed to offer little predictive power, with such infants generally having neurotypical development and similar 18 to 24-month cognitive, language, and motor outcomes as infants showing no visible injuries (***Wu et al., 2023; Bhroin et al., 2022***). Such claims, however, may stem from relying on visual perception to quantify signal abnormalities, or the use of a scoring system that underrepresents the contribution of injuries to structures like the amygdala and hippocampus (***Cizmeci et al., 2023***).

In this study, we created a population specific neonatal brain MRI template and obtained radiomic and geometric measures of the deep gray-matter structures and the brain as a whole. We explored how well these measures, and clinical and laboratory measures, predicted cognitive, language, and motor Bayley-III outcome scores. We used a machine learning method that could choose the best set of predictors from a large number of possibilities (Elastic-Net penalized linear regression), and found that the MRI-based measures yielded good predictions across all domains and across the full spectrum of outcomes. Indeed, with the MRI-based measures, the correlations between predicted and measured outcomes were more than twice that of the correlations for the predictions based on the clinical data alone, and explained between more than twice the variance in the observed outcomes. These findings show that quantitative neuroimaging measures can be effectively utilized with machine learning models to enhance the accuracy of neurodevelopmental outcome prediction in neonates with HIE across the full spectrum of outcomes.

In addition to demonstrating that brain MRI does have predictive power for the full spectrum of outcomes in neonates with HIE, our approach identified the structures and features within those structures that drove the predictions. This may provide valuable insight into our understanding of the outcomes associated with different patterns of injury in the developing brain. Notably, the top predictors of expressive language outcomes came from the amygdala, which suggests a surprisingly important role for the amygdala in this domain; and, all but one of the predictors were from the DGM structures, suggesting a lesser role for the circuitry associated with expressive language in adults, *e.g*. Broca’s area. Also of note, the hippocampus was one of the top predictors of receptive language outcomes. Injuries in the hippocampus have been associated with cognitive deficits (***Gadian et al., 2000***), however, to the best of our knowledge, have not been associated with receptive language deficits in the previous literature. As for motor outcome analyses, unsurprisingly, the top predictors included components of the DGM region but notably, features of the amygdala and hippocampus were also among the predictors, for gross motor and fine and composite motor, respectively.

Traditionally, the evaluation of brain injuries identified through MRI scans has been dependent on the subjective assessments made by radiologists. This method poses challenges related to consistency and reliability among different raters and even the same rater over time, leading to potential discrepancies and conflicting results. Moreover, certain brain injuries are so subtle that they might be overlooked or be undetectable by the human eye. We have addressed these issues by using radiomics measures that extract features directly from MRI data without human involvement. The features are derived from the size and shape of the labels, the image intensity histogram, the relationships between image voxels, gray-tone similarities and differences, and mathematically defined patterns, *e.g*. fractals. Most of these radiomics features are invisible to the human eye but computationally detectable. Furthermore, our use of an elastic-net model allowed for models that incorporate a large number of features but eliminated those features which do not contribute to the predictions. This method produces models that go beyond visual judgements of the severity of injury in different regions of the brain. The models produced by our iterative elastic-net regression approach found the best set of predictors for each structure, then combined them, and iterated to find the best combination of predictors, eliminating collinear predictors at each stage.

Our study has several limitations that should be considered when interpreting the results. First, a determination of the reliability of our results requires replication in a larger sample. It should also be noted that the predictive analyses were potentially negatively impacted by missing data in the clinical measures. The use of multiple imputations may have provided reasonable mitigation for this issue; however, we acknowledge this as a limitation. Second, our analyses may have also been negatively impacted by the use of different MRI scanners with different field strengths, even though scanner model was included as variable. Future studies should attempt to eliminate this variability in the data. The suboptimal slice thickness (3 mm, with 3.3 mm spacing) used in the protocol for the T2-weighted data might have also negatively impacted the analyses. This limitation necessitated acquiring good quality scans in all three orientations, using super-resolution up-sampling, and averaging to construct a T2-weighted scan of the same resolution as the T1-weighted scan. However, it should be noted that despite this limitation, the majority of the radiomics features that elastic-net used as predictors were taken from the T2-weighted data; thus, super-resolution up-sampling seems to produce reasonable results. Recent improvements to MR scanners allow the acquisition of the desired 3D T2-weighted scan directly and doing so would not only eliminate the need to perform this super-resolution up-sampling but may also yield superior results. Fourth, we were also limited by using only structural MRI data. Use of additional modalities, *e.g*. diffusion-weighted data, may allow for improved accuracy (***van Laerhoven et al., 2013; Thayyilet al., 2010***).

In conclusion, we have demonstrated that machine learning, using radiomics and geometric measures, has the potential to predict 18-month outcomes in infants with perinatal HIE across all domains, and across the full spectrum of outcomes. To contribute to the broader medical and neuroimaging communities, we provide our labeled multi-contrast population specific neonatal brain MRI template and the scripts necessary to use it to obtain the measures of the brain, and brain structures, and to produce the predictions of outcomes. However, we note that future studies with external cohort validation are needed to evaluate the generalizability of our findings.

## Data Availability

Our multi-contrast population specific neonatal brain MRI template and the labels for the deep gray-matter structures can be found here: https://gin.g-node.org/johndlewis/HIE/Template/  ; the scripts used to process the data can be found here: https://gin.g-node.org/johndlewis/HIE/Tools ; and the linear regression model can be found here: https://gin.g-node.org/johndlewis/HIE/Models/.

https://gin.g-node.org/johndlewis/HIE/Tools

https://gin.g-node.org/johndlewis/HIE/Models/

## Acknowledgments

Funding for this research was provided by the TD Bank Group Charity Classic Golf Tournament. Drs. Mehmet N. Cizmeci and Linh G. Ly received support from Dr. Karen Pape Program in Neuroplasticity for neuroprognostication and neurodevelopmental research. Our thanks to the Neonatal Neurodevelopmental Follow-Up Clinic team for their efforts to capture our outcome data. Also, our thanks to Vladimir S. Fonov and Jussi Tohka for their guidance.

## Author Contributions

JDL - conception and design; analysis of data; drafting the manuscript

AAM - data acquisition

MS - data acquisition HMB - data acquisition

AD - data acquisition

KR - data acquisition

LGL - data acquisition

MNC - conception and design; drafting the manuscript

BTK - conception and design; drafting the manuscript

## Data Availability

Our multi-contrast population specific neonatal brain MRI template and the labels for the deep gray-matter structures can be found here: https://gin.g-node.org/johndlewis/HIE/Template/ ; the scripts used to process the data can be found here: https://gin.g-node.org/johndlewis/HIE/Tools; and the linear regression model can be found here: https://gin.g-node.org/johndlewis/HIE/Models/.

